# Distinct components of cardiovascular health are linked with age-related differences in cognitive abilities

**DOI:** 10.1101/2022.02.22.22271137

**Authors:** Deborah L. O. King, Richard N. Henson, Rogier Kievit, Noham Wolpe, Carol Brayne, Lorraine K. Tyler, James B. Rowe, Cam-CAN, Kamen A. Tsvetanov

**Affiliations:** Department of Clinical Neurosciences, University of Cambridge, Cambridge CB2 0SP, United Kingdom; Centre for Speech, Language and the Brain, Department of Psychology, University of Cambridge, Cambridge CB23 6HT, United Kingdom; Department of Psychiatry, University of Cambridge CB2 2QQ, United Kingdom; Medical Research Council Cognition and Brain Sciences Unit, Cambridge CB2 7EF, United Kingdom; Cambridge Centre for Ageing and Neuroscience (Cam-CAN), University of Cambridge and MRC Cognition and Brain Sciences Unit, Cambridge CB2 7EF, United Kingdom; Donders Research Institute for Brain, Cognition and Behaviour, Radboud University, 6525 AJ Nijmegen, The Netherlands; Cambridge Public Health, Cambridge Public Health, University of Cambridge, Cambridge CB2 0SR, UK; Behavioural and Clinical Neuroscience Institute, Cambridge CB23 6HT, United Kingdom

## Abstract

Vascular, cardiovascular and neurovascular ageing processes contribute to cognitive impairment. However, the unique and synergistic contributions of cardiovascular factors to cognitive function remain unclear because they are often condensed into a single composite score or examined in isolation of each other. We hypothesized that vascular risk factors, electrocardiographic features and blood pressure indices reveal multiple latent vascular factors, with independent contributions to age differences in cognition. In a population-based deep-phenotyping study of cognition across the lifespan (n=708, age 18-88), path analysis revealed three latent vascular factors dissociating the autonomic nervous system response from two components of blood pressure. These three factors made unique and additive contributions to the variability in crystallized and fluid intelligence. The discrepancy in fluid relative to crystallized intelligence, indicative of cognitive decline, was associated with a latent vascular factor predominantly expressing pulse pressure, suggesting that higher pulse pressure is associated with cognitive decline from expected performance. This association varied with age, such that higher factor scores predicted a greater cognitive discrepancy in older individuals. Controlling pulse pressure may help to preserve cognitive function, particularly in older adults. Our findings highlight the need to better understand the multifactorial nature of vascular aging, its effects on neurocognitive function, and the modifiable risks of cognitive decline.

## INTRODUCTION

Life expectancy is increasing and the global population is ageing at an unprecedented rate. Identifying the factors that promote healthy cognitive ageing is therefore a public health priority (Brayne, 2007; Dreary et al., 2009), recognised by the World Health Organisation’s global strategy for collaborative action on healthy ageing (WHO, 2017). This includes identifying the risk and modifying factors for cognitive decline.

The second leading cause of cognitive decline in older people, after neurodegeneration, is vascular disease (Matthews et al., 2009), and vascular pathology is present in three-quarters of autopsies in older populations (Neuropathology Group, 2001). There may be a continuum between vascular pathology, dementia and Alzheimer’s Disease in the oldest old (Savva et al., 2009). Vascular factors trigger a cascade of cellular and molecular damage that remodels cerebral vessels and tissue (Barnes, 2015; de la Torre, 2012; Koenig et al., 2021; Qiu and Fratiglioni, 2015; Thorin-Trescases et al., 2018; Zimmerman et al., 2021). Vascular factors include total blood pressure (Jennings et al., 2021; Walker et al., 2017), pulse pressure (Waldstein et al., 2008; Mitchell et al., 2011; Meyer et al., 2017), heart rate variability (Appelhans and Luecken, 2006; Geisler et al., 2010; Koenig et al., 2021; Forte et al., 2019; Mather and Thayer 2018; Thayer et al., 2009, 2021; Varadhan et al., 2009) and body-mass index (Albanese et al., 2017; Kivimaki et al., 2018; Michaud et al., 2019; Veronese et al., 2017). Each factor may have different underlying causes and consequences for a spectrum of brain pathologies contributing to any degree of cognitive decline, ranging from subjective cognitive decline to dementia (van der Flier et al., 2018).

Ageing links vascular factors with cognitive decline, however the mechanisms underpinning this link are not well characterised. It is not established whether multiple vascular factors act synergistically through one shared biological pathway, or rather act independently with distinct – and possibly additive – effects on cognition. Vascular factors are often condensed into summary scores (DeRight et al., 2015; Veldsman et al., 2020), or considered in isolation from one another (eg Thayer et al., 2009). This approach hinders understanding of age-related changes in cognition, since different vascular factors may have different and interacting effects (Harada et al., 2013; Horn and Cattell, 1967; Shafto et al., 2020; Waldstein and Katzel, 2015). Furthermore, interactions of vascular factors with age, in predicting cognition, could be non-linear, but only linear effects are normally tested (e.g. Fuhrmann et al., 2019). Recent research indicates multiple, independent vascular pathways that are relevant to brain health and cognitive ageing (Fuhrmann et al., 2019; Qiu and Fratiglioni, 2015; Tsvetanov et al., 2021b). We propose that vascular ageing is better captured by multiple latent factors, and that these factors contribute differentially to age-related changes in cognitive abilities.

Intelligence is a complex construct, in which crystallized and fluid intelligence are often distinguished (Cattell, 1963). Crystallized intelligence represents acquired and general knowledge, remains relatively stable throughout life, and can be used to index baseline intelligence (Deary and Caryl, 1997). Fluid intelligence represents flexible problem-solving ability, encompassing working memory and executive functions, and typically declines steadily from early adulthood (Manard et al., 2014; de Mooij et al., 2018; Shafto et al., 2020). Though they are positively correlated across individuals, the difference between crystallized and fluid intelligence – the “discrepancy” – is a sensitive measure of within-subject decline arising from brain injury, degeneration and ageing (Dierckx et al., 2008; Harrington et al., 2018; McCarthy et al., 2005; McDonough et al., 2016; McDonough and Popp, 2020; O’Carrol and Gilleard, 1986; O’Shea et al., 2018; Rabbit, 1993; Tomassini et al., 2021). A higher discrepancy score, when subtracting normalised fluid intelligence from normalised crystallized intelligence, indicates greater cognitive decline (McDonough et al., 2016). Little is known about what determines the degree of discrepancy in older age. However, vascular and degenerative factors are proposed to contribute. For example, the ability discrepancy has been associated with amyloid beta deposition (McDonough et al., 2016). Despite the value of such an estimate of decline, one can also measure diverse functions to better characterise the complex construct of cognition.

The present study investigated the relationship between multiple vascular measures and the cognitive ability discrepancy from the Cambridge Centre for Ageing and Neuroscience (Cam-CAN; www.cam-can.org), with 708 adults, aged 18 to 88 (Shafto et al., 2014). The vascular measures were body mass index (BMI), heart rate, heart rate variability (represented by both low and high frequencies of the electrocardiogram, ECG (Forte et al., 2019), and blood pressure. Instead of representing blood pressure through simple systolic and diastolic measures, better insight may be achieved by transforming it into its steady and pulsatile components (Darne et al., 1989; Kim et al., 2011; Laosiripisan et al., 2017; Lefferts et al., 2016; Roman and Devereux, 2014; Smulyan and Safar, 1997; Strandberg and Pitkala, 2003; Tanaka et al., 2016). Therefore, we report pulse pressure (difference between systolic and diastolic blood pressure) and total blood pressure (the sum of systolic and diastolic, to be orthogonal to pulse pressure). The observed vascular variables were then modelled with exploratory factor analysis, with the expectation of three latent vascular factors based on previous work (Tsvetanov et al., 2021b). The observed cognitive measures were the Spot-The-Word and Proverbs tests, believed to capture crystallized intelligence, and the four sub-scores on the Cattell test, believed to capture fluid intelligence (see Shafto et al., 2014, for details). A confirmatory factor analysis was used to define the two Latent Cognitive Factors (LCF) of fluid and crystallized intelligence. The difference between these LCFs produced the ability discrepancy score (McDonough et al., 2016). The relationships between the ability discrepancy, the three latent vascular factors and age, as well as their interactions, were then investigated with multiple linear regression. We examined interactions with sex and medication (binary for a number of drugs relevant to cardiovascular and cognitive health) (Marvanova et al., 2016; Schultz et al., 2018; Yang et al., 2020). We covaried education level, which may capture baseline differences in fluid intelligence not represented by our measures of current crystallized intelligence (Rönnlund et al., 2005).

We predicted (i) that latent Vascular Factors associated with “good” cardiovascular health would decrease with age, while the cognitive ability discrepancy would increase with age; and (ii) that some latent Vascular Factors would associate with the ability discrepancy over and above age, and with a strength of association that changes with age, whereby ability discrepancy in older people would be more dependent on their latent vascular factors scores.

## METHODS

### Participants

Figure 1 illustrates the analytical strategy and the study design with the Cam-CAN cohort (n=708, Shafto et al., 2014; Taylor et al., 2017). Ethical approval was obtained from the Cambridgeshire 2 (now East of England—Cambridge Central) Research Ethics Committee. Participants were recruited from Cambridge City GPs, randomly selected from this complete population sampling frame. Participants gave written informed consent. The detailed recruitment pathway is outlined in Supplementary Figure 1. Exclusion criteria are described in full detail elsewhere (Shafto et al., 2014); in brief, those participating represented the healthier and more advantaged spectrum within the population at all ages. The diastolic and systolic blood pressure observations were excluded for one participant due to data entry errors. Education was reported across four categories of: none, GCSE or O-Level, A-Level, Degree (College or University). Medication status (binary on/off) was reported across four categories of drugs with cardiovascular relevance: (1) anti-hypertensives (Yang et al., 2020), including angiotensin receptor blockers, angiotensin converting enzymes inhibitors, calcium channel blocking agents and thiazide diuretics; (2) beta blockers, including beta selective and non-selective beta blockers; (3) other diuretics, including loop and potassium sparing diuretics; (4) dyslipidemics drugs, including statins (Marvanova et al., 2016; Schultz et al., 2018; Yang et al., 2020).

**Figure 1.**
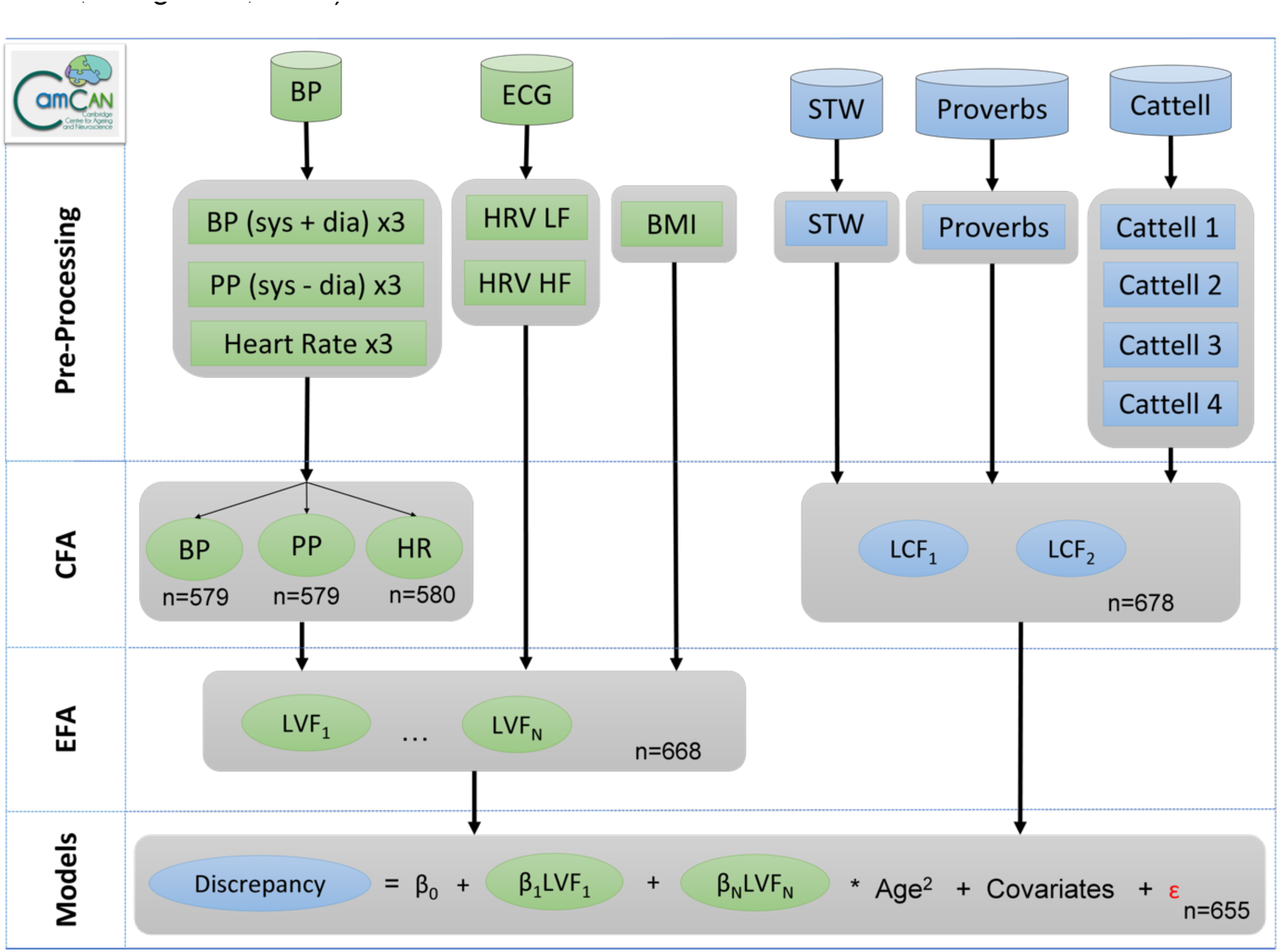
Schematic representation of the data processing and analysis pipeline to investigate shared and unique relationships between vascular and cognitive factors in the Cam-CAN dataset (n=708). Abbreviations: BMI, body mass index; BP, total blood pressure (systolic + diastolic); dia, diastolic; Cattell 1-4, sub-scores across the four Cattell tasks; CFA, confirmatory factor analysis; Discrepancy, the ability discrepancy, defined as LCF2 (crystallized) minus LCF1 (fluid); ECG, electrocardiogram; EFA, exploratory factor analysis; HR, heart rate; HRV HF, heart rate variability at high frequency; HRV LF, heart rate variability at low frequency; LCF, latent cognitive factor; LVF, latent vascular factor; PP, pulse pressure (systolic – diastolic); sys, systolic; STW, spot the word.

### Vascular Factors

Systolic and diastolic blood pressure, and heart rate, were measured using the A&D Medical Digital Blood Pressure Monitor (UA-774). Measurements were taken after at least 10 minutes of a participant being seated and repeated 3 times in succession. BMI was calculated using portable scales as weight (kg)/height (m)^2^. Heart rate variability was based on the frequency-domain information of normal-to-normal beats and extracted from resting state electrocardiogram recordings while seated during a separate MEG scan. We separated low-and high-frequency components: high frequency heart rate variability (0.15–0.4 Hz) principally indexes parasympathetic vagal influences, while low frequency heart rate variability (0.05–0.15 Hz) indexes non-vagal and sympathetic nervous system influences (Forte et al., 2019; Laborder et al., 2017). These two branches of the autonomic nervous system exhibit different non-linear trajectories with age, and might relate differently to cognition (Zulfiqar et al., 2010). Heart rate variability data was processed using the PhysioNet Cardiovascular Signal Toolbox (Goldberger et al., 2000; Vest et al., 2018) in Matlab (MATLAB 2017b, The MathWorks Inc, Natick, MA). Following Tsvetanov et al. (2021b), segments classified as atrial fibrillation were excluded and data for any participant with >50% atrial fibrillation (n=1) were excluded. The heart rate variability at low and high frequency, and BMI, were log-transformed to render them more Gaussian.

### Behavioural tasks

Crystallized intelligence was assessed through the Spot the Word and Proverb Comprehension tasks. In the Spot the Word test of vocabulary, participants were asked to point to the letter string in a pair that is a real word (albeit infrequent) (Baddeley et al., 1993). In Proverb Comprehension, participants read and interpreted three English proverbs (Hodges, 1994). Fluid intelligence was assessed with the Cattell Culture Fair Test, in which participants completed non-verbal puzzles resulting in four summary scores based on series completion, classification, matrices and topology conditions (Cattell, 1971; Kievit et al., 2014).

### Statistical Analyses

Analyses were performed in R-Studio (R version 4.0.1, 2019; R Core Team, 2020). For initial checks, the three observations of diastolic and systolic pressure were correlated with age, using the Pearson’s product moment correlation coefficient. They were then used to calculate three sets of total blood pressure (systolic + diastolic) and pulse pressure (systolic – diastolic). The resulting scores, and the three observations of heart rate, were log transformed to conform more closely to Gaussian distributions. These three sets of blood pressure, pulse pressure and heart rate measures were then standardised (mean=0, standard deviation=1) and condensed into a single latent variable per domain, using confirmatory factor analysis (CFA) in the Lavaan package (Rosseel, 2012). Latent variables reduce error and estimation bias, while increasing precision (Little et al., 2002, 1999). Note that the confirmatory models were saturated, where three indicators loading onto one latent variable gave zero degrees of freedom. Missing data were imputed using Full Information Maximum Likelihood in cases where data were recorded for at least one of the three domain-specific observations; where data were missing entirely, participants were omitted automatically (leaving n=579 for blood pressure and pulse pressure; n=580 for heart rate).

Next, we sought to identify the optimal number of factors among all vascular variables using Exploratory Factor Analyses (EFA). EFA is a common multivariate statistical method used to uncover the structure of a large set of variables (Watkins, 2018). Thus, EFA was used to identify the smallest number of latent vascular factors (latent vascular factors) that can parsimoniously explain the covariance observed among all vascular variables (blood pressure, n=579; pulse pressure, n=579; heart rate, n=580; heart rate variability, at low and high frequencies, n=604; BMI, n=587). EFA was performed with the Psych package and imputing missing data (n=668) (Revelle, 2017). Allowing two variables per latent factor is the upper boundary limit for model identification, meaning that 1-, 2-and 3-factor solutions can be explored for six vascular variables in the current study. Note that a 3-factor model with 6 variables will be fully saturated, not allowing estimation of the absolute fit indices. Therefore, model validity was based on model comparisons using the Chi-squared statistic, i.e. using comparative fit indices to determine the optimal number of latent vascular factors. To further explore the robustness of the winning model (and its loadings), we performed an additional structural equation model that included cognitive variables too (see Supplementary Section B). Factor score estimates for each latent vascular factor were then extracted from the winning model for further regression analysis below.

Observed cognitive variables were standardised and condensed into two latent variables, using confirmatory factor analysis. The two-factor structure was based on the established dissociation between crystallized and fluid intelligence (Cattell, 1943). Scores on the Proverbs (n=655) and Spot the Word tests (n=705) loaded onto one latent cognitive factor (LCF1), representing crystallized intelligence. Scores on the Cattell tests (n=660) loaded onto LCF2, representing fluid intelligence. Missing data were imputed using Full Information Maximum Likelihood in cases where data were recorded for at least one observed variable, producing LCFs for n=678. The difference between LCF1 and LCF2 was calculated to give the ability discrepancy (McDonough et al., 2016).

The latent vascular factors and ability discrepancy were standardised to allow interpretation in terms of standard deviations from the mean. Linear and quadratic age predictor terms were also standardised. The relationships between the latent vascular factors, ability discrepancy and age were examined in multiple linear regression, using complete case analysis (n=655). The presence of outliers with undue influence, as identified with Cook’s criteria (Cook, 1997), motivated the use of robust linear regression, implemented in the Robustbase package (Maechler et al., 2021). We performed a series of regression models from simple to complex, all including sex and education as covariates of no interest, but dropping effects that did not improve overall model fit. Fit was investigated with the Akaike Information Criterion, Bayesian Information Criterion and Sum of Squares. Results were reported at p<0.05. To guide the interpretation of significance of parameters in the non-winning models, model specific p-values after Bonferroni corrections are also reported.

Five models were used to test different hypotheses. The first model examined the relationship between the ability discrepancy and the three latent vascular factors, ignoring any shared dependence on age, to reveal which latent vascular factor (s) make unique contributions to the ability discrepancy. In the following model syntax, latent vascular factors are abbreviated to LVF1-3.

Model 1:

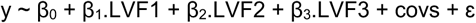

where “y” is ability discrepancy and “covs” are the covariates of no interest, i.e, sex and education, “β” are the parameter estimates (coefficients) and “ε” is the residual error.

The Second model investigated whether any relationships between the ability discrepancy and latent vascular factors remained over and above a second-order polynomial expansion of age, and/or whether any effects of latent vascular factors depended on age. Note that, since the latent vascular factors were highly correlated with age, if effects of latent vascular factors from Model 1 are no longer significant in Model 2, then this could simply be because age shares variability with the latent vascular factors.

Model 2:

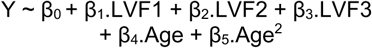

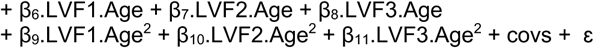

Models 1 and 2 were compared and if Model 2 was shown to better fit the data, then the age terms were taken forwards into further models.

Model 3 investigated whether our findings could be explained by medication status.

Model 3:

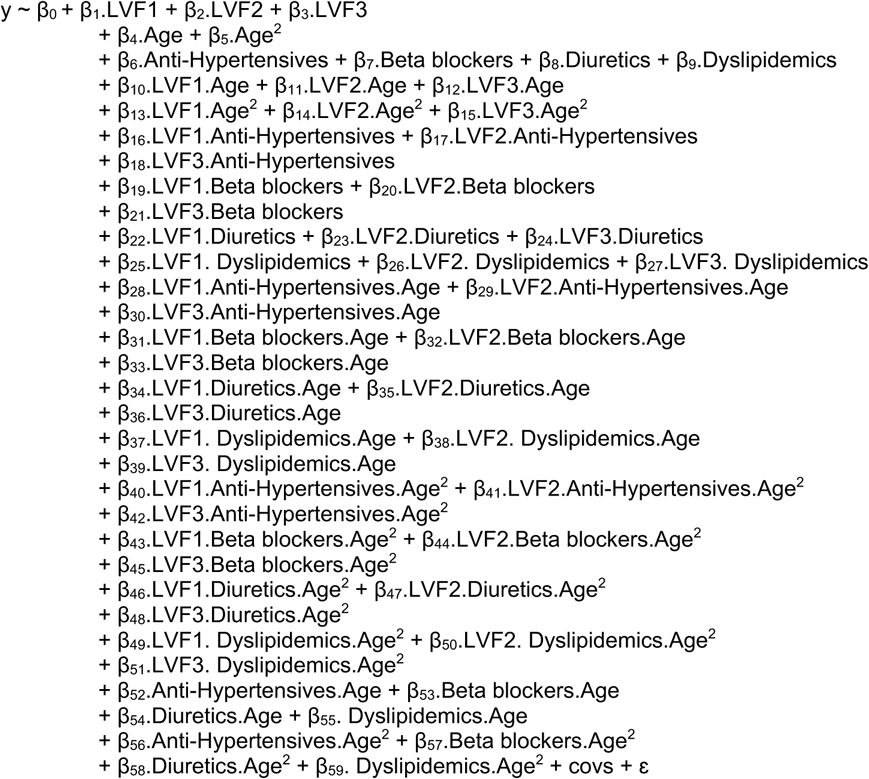

Model 4 accounted for the interacting effects of sex on Vascular factors with age (Klassen et al., 2021; Koenig et al., 2021; Reas et al., 2021; Thayer et al., 2021). Since model comparisons showed that medications did not improve overall fit in Model 3, medications were not specified in Model 4.

Model 4:

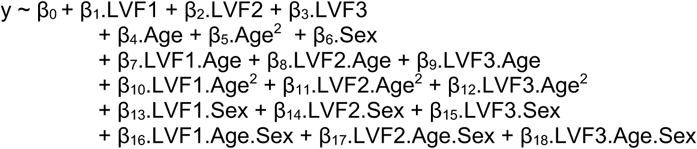

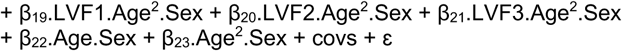

Model 5 investigated whether latent vascular factors interact with each other in order to determine the ability discrepancy. Since model comparisons showed that the inclusion of Sex interaction terms did not improve overall fit in Model 4, these interactions were not perpetuated to Model 5.

Model 5:

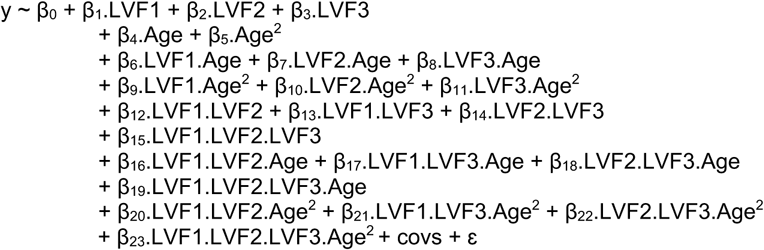

## RESULTS

### Participants

Characteristics of the 708 participants in the CamCAN Phase 2 are outlined in Table 1. Rate of missing data varied between 0 and 18% (see Table 1). When the cohort (n=708) was split into three age groups, the proportion of each on medications was: 0% of the younger (18-37 years); 12.6% of the middle (38-67 years) and 51.6% of the older (68-88 years) group. Across the entire cohort, 6.5% of participants had no education beyond 15 years; 13.6% had GCSEs or O-levels; 19.5% had A-levels; and 60.2% had a degree.

**Table 1.** Demographic information by age-tertiles (n=708). For the three observations of systolic, diastolic and heart rate, average values are reported. Measures presented here were averaged over the three observations. One decimal place is reported where data are continuous.

|  | Complete Data (n) |  |  | Missing (%) |  |  | Range / Number (Male) |  |  | Mean |  |  | SD |  |  |
| --- | --- | --- | --- | --- | --- | --- | --- | --- | --- | --- | --- | --- | --- | --- | --- |
|  | Young | Middle | Old | Young | Middle | Old | Young | Middle | Old | Young | Middle | Old | Young | Middle | Old |
| Age (years) | 164 | 325 | 219 | 0 | 0 | 0 | 18-37 | 38-67 | 68-99 | 29.5 | 52.5 | 76.6 | 5.5 | 8.5 | 5.4 |
| Sex (Male) | 164 | 325 | 219 | 0 | 0 | 0 | 79 | 159 | 111 | - | - | - | - | - | - |
| Diastolic (mmHG) | 142 | 265 | 169 | 13.4 | 18.5 | 22.8 | 51.7-94.3 | 50-118.7 | 49-114.3 | 69.9 | 74.9 | 72.4 | 8.7 | 10.4 | 10.7 |
| Systolic (mmHG) | 142 | 265 | 169 | 13.4 | 18.5 | 22.8 | 92.3-141 | 79.3-172.3 | 82.3-178.3 | 112.3 | 118.7 | 130.1 | 11 | 14.6 | 18.9 |
| Heart Rate (beats/minute) | 142 | 265 | 169 | 13.4 | 18.5 | 22.8 | 43.3-95.7 | 39-96.3 | 44.7-107.7 | 65.5 | 64.5 | 68 | 10.1 | 9.5 | 12 |
| HRV low frequency (ms<sup>2</sup>) | 135 | 299 | 170 | 17.7 | 8 | 22.4 | 5.5-9283.7 | 13-15784.9 | 1.3-1304.4 | 1163 | 834.9 | 231.8 | 1184.3 | 1393.2 | 257.7 |
| HRV high frequency (ms<sup>2</sup>) | 135 | 299 | 170 | 17.7 | 8 | 22.4 | 23.4-7129.8 | 10.2-5332.9 | 5.5-3543.8 | 1422.3 | 648.2 | 264.6 | 1226.6 | 767.3 | 378.1 |
| BMI (kg/m<sup>2</sup>) | 142 | 268 | 177 | 13.4 | 17.5 | 19.2 | 16.8-37.7 | 17.6-48.3 | 19.9-44.3 | 23.8 | 25.9 | 27.1 | 4 | 4.9 | 4 |
| Cattell sub-score 1 | 154 | 307 | 199 | 6.1 | 5.5 | 9.1 | 6-12 | 3-12 | 2-12 | 10.6 | 9.8 | 7.8 | 1.3 | 1.7 | 2.1 |
| Cattell sub-score 2 | 154 | 307 | 199 | 6.1 | 5.5 | 9.1 | 3-13 | 3-13 | 1-12 | 9.6 | 8.3 | 6.7 | 1.9 | 1.9 | 1.9 |
| Cattell sub-score 3 | 154 | 307 | 199 | 6.1 | 5.5 | 9.1 | 7-12 | 4-12 | 1-12 | 10.6 | 9.5 | 7.2 | 1.3 | 1.7 | 2 |
| Cattell sub-score 4 | 154 | 307 | 199 | 6.1 | 5.5 | 9.1 | 1 to 8 | 1 to 9 | 1 to 8 | 6.4 | 5.4 | 4.1 | 1.4 | 1.6 | 1.9 |
| Proverbs | 149 | 310 | 196 | 9.1 | 4.6 | 10.5 | 0-6 | 0-6 | 0-6 | 4.1 | 4.7 | 4.6 | 1.7 | 1.6 | 1.7 |
| Spot the Word | 163 | 325 | 217 | 0.6 | 0 | 0.9 | 24-60 | 30-60 | 29-60 | 51.4 | 54.2 | 54.3 | 5.8 | 4.5 | 5.9 |
| Medications<br>(percentage of total per drug) | - | - | - | 0 | 0 | 0 | - | - | - | - | - | - | - | - | - |
| Anti-hypertensives | 0.0 | 8.0 | 37.4 | - | - | - | - | - | - | - | - | - | - | - | - |
| Beta Blockers | 0.0 | 0.9 | 9.6 | - | - | - | - | - | - | - | - | - | - | - | - |
| Other diuretics | 0.0 | 2.8 | 13.2 | - | - | - | - | - | - | - | - | - | - | - | - |
| Dyslipidemics | 0.0 | 8.6 | 28.3 | - | - | - | - | - | - | - | - | - | - | - | - |
| Education<br>(percentage of total by category) | - | - | - | 0.6 | 0.0 | 0.5 | - | - | - | - | - | - | - | - | - |
| No qualifications tried (< 16) | 0.6 | 3.1 | 16.0 | - | - | - | - | - | - | - | - | - | - | - | - |
| GCSEs / O-levels (age 16) | 11.0 | 14.2 | 14.6 | - | - | - | - | - | - | - | - | - | - | - | - |
| A-levels (age 18) | 15.2 | 18.5 | 24.2 | - | - | - | - | - | - | - | - | - | - | - | - |
| Degree (over 18) | 72.6 | 64.3 | 44.7 | - | - | - | - | - | - | - | - | - | - | - | - |

### Vascular Factors and Age

Diastolic and systolic blood pressure increased with age (Supplementary Figure 2). Within each confirmatory factor analysis on the repeated measures of blood pressure, heart rate and pulse pressure, there were positive associations between all observed and latent variables, with p<0.001 for all factor loadings. The resulting latent variables for blood pressure, heart rate and pulse pressure therefore showed significant positive associations with age (Supplementary Figure 3). BMI also correlated significantly positively with age, while heart rate variability at both frequencies showed a significantly negative association (Supplementary Figure 3).

### Vascular Factor Analysis

We used EFA to estimate models with one, two and three factors. The two-factor model did not converge well. The three-factor model was fully saturated (no residual degrees of freedom). Model comparisons showed two-factors to fit better than one (Δ *X^2^* = 236.57, p<0.001), and three-factors to fit better than two factors (Δ *X^2^* = 95.04, p<0.001 – but note the limitation that the two-factor solution was improper). The estimates of factor loadings and covariances is visualised in Figure 2A and detailed fully in Supplementary Table 1. All latent vascular factors correlated significantly with age (Figure 3). We confirmed the validity of the three-factor model in combination with cognitive measurements (see Supplementary Section B).

**Figure 2.**
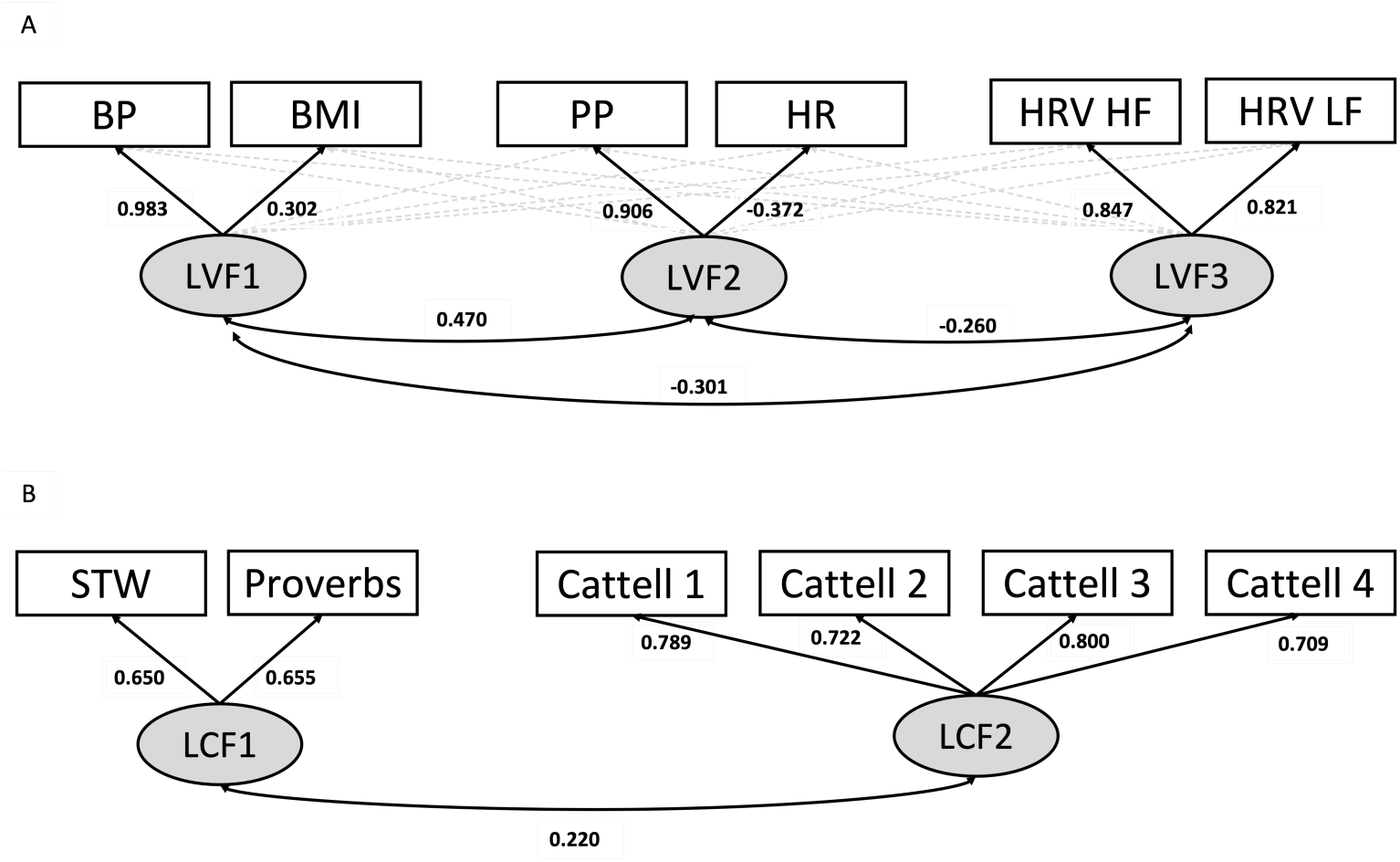
A) The three-factor Exploratory Factor Analysis model of vascular health. The numeric values of cross-loadings <0.30 (dashed grey arrows) are omitted here for visual clarify and reported fully in Supplementary Table 1. B) The two-factor Confirmatory Factor Analysis model of cognition. Abbreviations: BMI, body mass index; BP, total blood pressure; HR, heart rate; HRV HF, heart rate variability at high frequency; HRV LF, heart rate variability at low frequency; LCF, latent cognitive factor; LVF, latent vascular factor; PP, pulse pressure; STW, spot the word.

**Figure 3.**
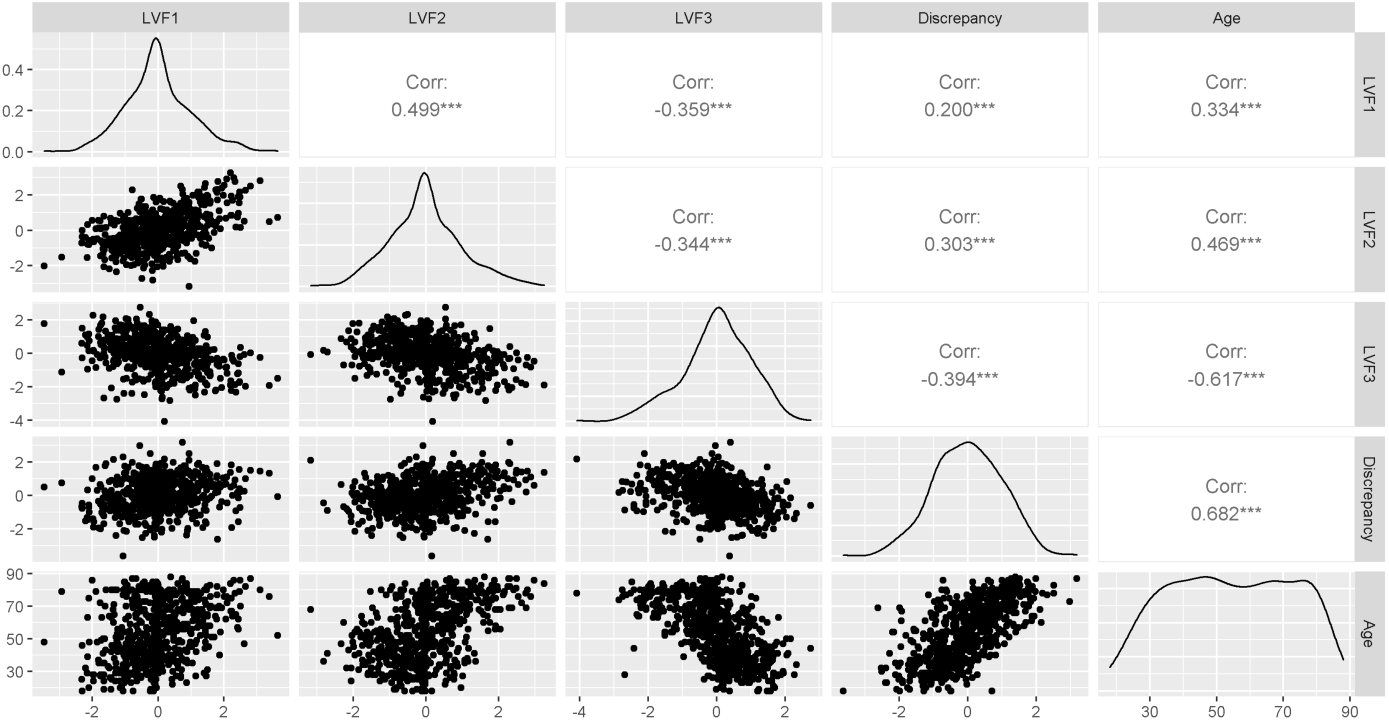
Scatter plots (lower left), distributions (leading diagonal) and Pearson correlations (upper right) for Latent vascular factors, ability discrepancy and age. Stars indicate increasing significance on the correlations: ***, p<0.001; **, p<0.01; *, p<0.05. Abbreviations: Corr, correlation coefficient; Discrepancy, ability discrepancy; LVF1-3, latent vascular factors 1-3.

### Ability Discrepancy

The cognitive variables (Supplementary Figure 4) were entered into a two-factor CFA and produced latent cognitive factor 1, representing crystallized intelligence, and latent cognitive factor 2, representing fluid intelligence (Figure 2B). The ability discrepancy was calculated as latent cognitive factors 1 – 2 (McDonough et al., 2016). The ability discrepancy showed a strong positive association with age (Figure 3). It also correlated significantly with the three latent vascular factors, with substantial effect sizes (Figure 3).

### Multiple Linear Regression

In Model 1(n=655, R^2^=0.220, adjusted R^2^=0.210), ability discrepancy showed a significant positive relationship with latent vascular factor 2 (std β=0.212, SE=0.043, p<0.001) and a significant negative relationship with latent vascular factor 3 (std β=-0.339, SE=0.039, p<0.001) (Supplementary Table 2). Thus, while all three latent vascular factors explained shared variance, only latent vascular factors 2 and 3, but not 1, made unique contributions to the ability discrepancy.

Compared to Model 1, Model 2 (n=655, R^2^=0.511, adjusted R^2^=0.498) fit the data better (Supplementary Table 3). Model 2 showed that the main effects of latent vascular factors 2 and 3 did not remain significant when accounting for age. This is likely explained in part by their shared variance with age (for which the linear effect was significant, std β=0.713, SE=0.046, p<0.001). More interesting was a significant interaction between latent vascular factor 2 and the quadratic effects of age (std β=0.087, SE=0.037, p=0.018). This interaction is visualised in Figure 4, by splitting the data into three age groups. It can be seen that the positive relationship between latent vascular factor 2 and the ability discrepancy is over 7 times stronger in the older group compared with the two younger groups.

**Figure 4.**
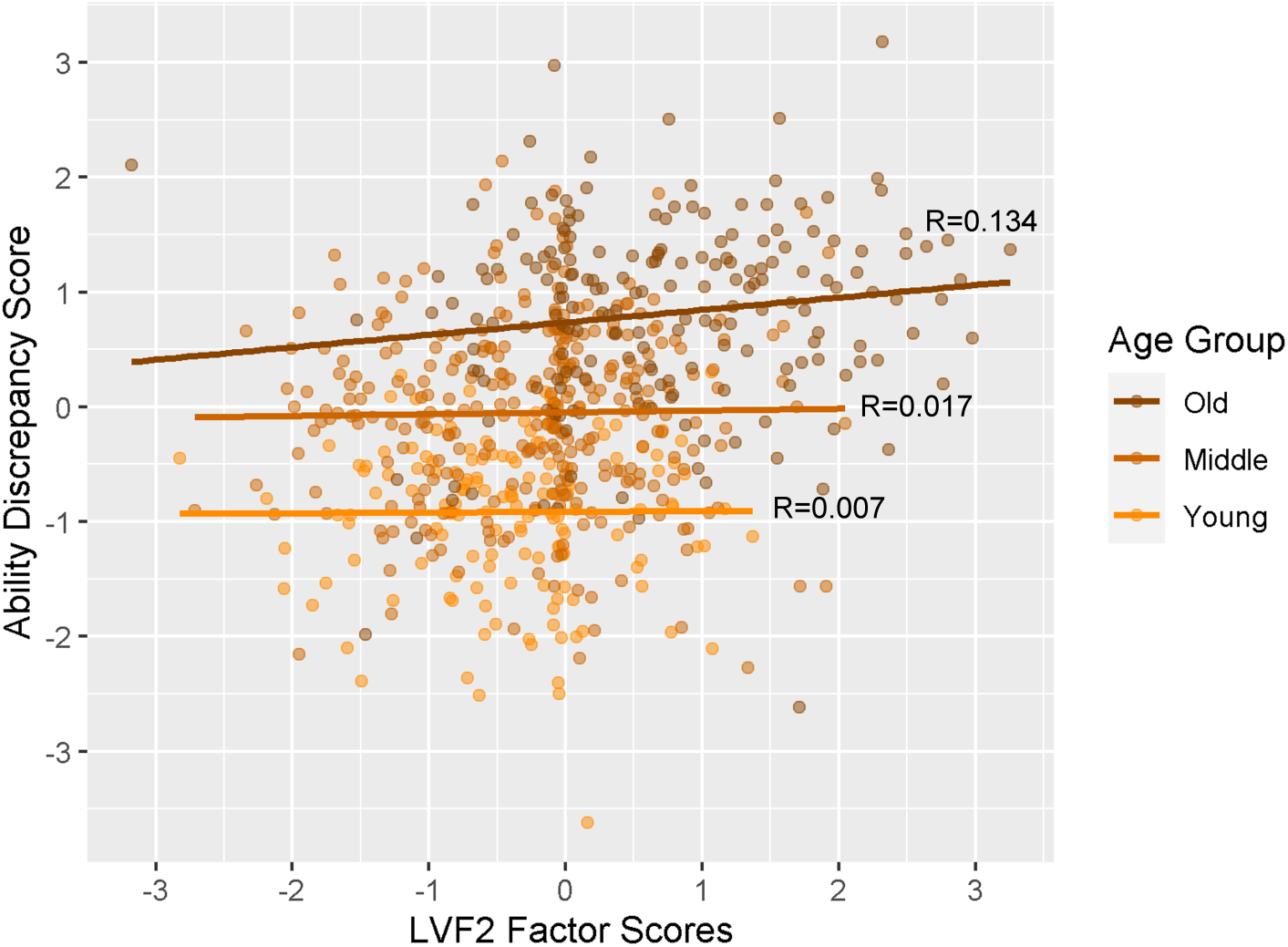
A visualisation of the effect of latent vascular factor 2, expressing predominantly pulse pressure, on the ability discrepancy for complete case data (n=655). Note that age was a continuous variable for the interaction tested, but here participants are plotted as discrete groups of young (18-37 years, n=154), middle (38-67 years, n=307) and old (68-88 years, n=194) for visualisation purposes only. Abbreviations: LVF2, latent vascular factor 2.

From Model 2, there was no significant improvement in fit when adding medications (Model 3), interactions with sex (Model 4), or interactions between Latent vascular factors (Model 5) (Supplementary Table 4). Given the lack of evidence supporting these more complex models, any significant parameters in Models 3-5 (Supplementary Tables 5-7) should be considered as suggestive only. Furthermore, this significance may not survive correction for multiple comparisons.

## DISCUSSION

Here we confirm that changes in vascular systems across the lifespan have multifactorial effects on cognitive function. There are three key observations. First, we identify three latent vascular factors that broadly dissociate the autonomic nervous system from distinct components of blood pressure. Second, these factors make distinct contributions to age-related cognitive decline, as indexed by the “ability discrepancy” score (McDonough et al., 2016). Third, the pulse pressure was particularly associated with the cognitive discrepancy, increasingly so for older adults. This remained even after controlling for the use of hypertensive medications and the covariates of sex and education. Importantly, the effect of pulse pressure was independent of other latent vascular factors. We propose that steps to maintain lower pulse pressure may help to preserve cognitive function into old age.

### Three components of Vascular Health

A single composite measure was insufficient to capture vascular health in our exploratory analyses, and the model fit was better with three factors. The evidence for these multiple latent vascular factors is consistent with previous model-based and data-driven approaches (Chen et al., 2000; Fuhrmann et al., 2019; Goodman et al., 2005; Khader et al., 2011; Mayer-Davis et al., 2009; Tsvetanov et al., 2021b). The three latent vascular factors were composed predominantly of two major blood pressure variables and an autonomic nervous system variable; a decomposition that agrees with previous studies (Mayer-Davis et al., 2009; Tsvetanov et al., 2021b). This decomposition also mimics established models of cardiovascular health (discussed below). The unsupervised construction of these latent vascular factors highlights their distinct contributions, which may involve different pathways and require different interventions.

The first factor, latent vascular factor 1, expressed total blood pressure, which can be considered a steady state component of blood pressure (Figure 2). This component is proposed to be mainly influenced by cardiac output and peripheral vascular resistance (Smulyan and Safar, 1997). The additional contribution of BMI to latent vascular factor 1 fits well with early work showing a strong correlation between BMI and the steady component of blood pressure (Darne et al., 1989). Latent vascular factor 1 was positively associated with age and the ability discrepancy (Figure 3), however it did not significantly predict the ability discrepancy, over and above other latent vascular factors, in Model 1 (Supplementary Table 2). Consistent with these observations, Lefferts et al. (2016) showed that steady blood pressure no longer predicts cognition over and above the effects of pulsatile blood pressure and covariates (Lefferts et al., 2016). We previously found that the steady component of blood pressure is associated with age-related cerebrovascular dysfunction of the sensorimotor regions, independently of the pulsatile component (Tsvetanov et al., 2021b). This suggests a unique contribution of steady blood pressure to brain health, with regional specificity. Future work should establish the possibility of a specific contribution of steady state blood pressure to brain functioning, and whether this varies across the lifespan (Wills et al., 2011).

The second component, latent vascular factor 2, expressed pulse pressure, with some contribution from heart rate (Figure 2). Latent vascular factor 2 was positively associated with age (Figure 3). Given the predominant loading by pulse pressure, latent vascular factor 2 likely represents a cerebrovascular element (Levin et al., 2020; Wåhlin and Nyberg, 2019). This is also consistent with previous findings that pulse pressure and white matter lesion burden expressed a common latent cerebrovascular factor (Tsvetanov et al., 2021b).

Latent vascular factor 3 expressed resting heat rate variability (Figure 2), which indexes a specific component of the autonomic nervous system (Malik et al., 1996; Porges, 2003; Thayer, 2009). The decomposition of autonomic nervous system signals separately from cerebrovascular health signals (latent vascular factors 1 and 2) confirms that these are distinct, yet partly correlated, constructs of vascular ageing (Schroeder et al., 2003). The convergence of low and high frequencies of resting heart rate variability onto latent vascular factor 3 does not necessarily rule out frequency-specific effects on cognition, or frequency-specific effects of task-based heart rate variability modulation/reactivity (Laborde et al., 2018; Lui et al., 2022). Latent vascular factor 3 associated negatively with age, consistent with previous studies on heart rate variability (Choi et al., 2020).

### Pulse Pressure and age-related Cognitive function

A novel aspect of our work was to simultaneously relate the three latent vascular factors to age-related differences in cognition, specifically the cognitive discrepancy score. In the absence of longitudinal data, this discrepancy is a better estimate age-related change than (baseline) individual differences. When relating directly to cognitive ability, only latent vascular factors 2 and 3 made unique contributions. Note that this does not mean latent vascular factor 1 has no relationship with cognitive ability; only that we cannot distinguish any such contribution from those of factors 2 and 3. The negative relationship for latent vascular factor 3 shows that higher heart-rate variability is associated with lower ability discrepancy, i.e., more variable heart rate is associated with less discrepancy, i.e., fluid intelligence that is closer to what would be expected from crystallized intelligence.

However, the relationship between latent vascular factor 3 and cognitive discrepancy was no longer significant when age was controlled for. This is consistent with cross-sectional studies where the association between resting heart rate variability and executive functions is accounted for by age and systemic vascular health (Kimhy et al., 2013; Mann et al., 2015; Stenfors et al., 2016). Future studies should investigate how the shared variance between heart rate variability, systemic vascular health, age and cognition is linked to changes in cerebral blood flow, tissue integrity and neural function (Forte et al., 2019; Koenig et al., 2021; Shaffer and Ginsberg, 2017). Heart rate variability is also theorised to link to domain specific measures of cognition, including emotional regulation (Forte et al., 2019; Holzman and Bridgett, 2017; Mather and Thayer 2018; Thayer et al., 2009, 2021). Future research should also explore whether heart rate variability affects emotional regulation independently to other Vascular factors, and whether this relationship changes with age.

Latent vascular factor 2 made a unique, positive contribution to ability discrepancy, consistent with higher pulse pressure being detrimental to cognitive ability. Like latent vascular factor 3, this contribution was no longer significant when adding age to the model. This could be because age is the true driver of ability discrepancy, or that age and latent vascular factor 2 are so highly correlated that we can no longer detect a unique effect of LVF2. More importantly, we did find a significant interaction between age and latent vascular factor 2. This was a quadratic effect, consistent with pulse pressure being especially important for cognition in old age, rather than changing linearly with age. These findings are consistent with a growing body of literature suggesting that pulsatile, rather than steady, blood pressure is an important factor for brain health and higher cognitive functions (Lefferts et al., 2016; Mitchell, 2008; Mitchell et al., 2011; Obisesan et al., 2008; Waldstein et al., 2008; Webb et al., 2012). Future work needs to evaluate whether maintaining normal pulse pressure across the lifespan is the mediating factor of cognitive function and play a role in the increasing relationship between brain function and cognition in old age (Bethlethem et al., 2020; Tibon et al., 2021; Tsvetanov et al., 2016, 2018; 2021a).

The mechanism by which pulse pressure relates to the ability discrepancy has yet to be identified. Pulse pressure has been proposed to be associated with a trigger point of a positive feedback loop of rising arterial stiffness and pressure that penetrates increasingly into deep brain tissue (Thorin-Trescases et al., 2018). This causes a cascade of molecular and cellular damage to cerebral vessels, which ultimately injures the blood brain barrier, promoting the aggregation of beta-amyloid (Hughes et al., 2013; Rodrigue et al., 2013; Stone et al., 2015), phosphorylated tau (Nation et al., 2015) and white matter hyperintensities (Levin et al., 2020; de Montgolfier et al., 2019; Thorin-Trescases et al., 2018; Wåhlin and Nyberg, 2019). Pulse pressure induced hippocampal damage is theorised to result in impaired episodic memory (Wåhlin and Nyberg, 2019), while it also uniquely contributes to cerebrovascular dysfunction in frontoparietal regions (Tsvetanov et al., 2021b). The recruitment of frontoparietal regions is of particular interest here, given that their involvement in fluid ability processing is partly explained by age-related hypoperfusion in these areas (Wu et al., in press). Separately, pulse pressure is associated with amyloid-dependent hypometabolism in frontoparietal regions (Langbaum et al., 2021), which has in turn been associated with the ability discrepancy score (McDonough et al., 2016). Our research adds to this evidence, by showing that pulsatile, but rather than steady state blood pressure, is a predictor of ability discrepancy, This effect is greater so for older adults, in a population-based lifespan cohort. Future research should investigate whether pulse pressure affects the ability discrepancy and episodic memory through shared or separate mechanisms. We propose that pulse pressure links to cognitive ageing through a distinct mechanism of cellular and molecular changes in cerebral vessels (Wåhlin and Nyberg, 2019; Tsvetanov et al., 2021b; Wu et al., in press). We further highlight pulse pressure as an emerging therapeutic target to prevent cognitive decline in ageing (Levin et al., 2020).

### Limitations

Our study has limitations. The results were based on a population-based cross-sectional cohort and cannot directly speak to dynamic ageing, that is to say individual subjects’ progression over time. Thus, our discussion of age effects was restricted to the effects of age and its correlates, as assessed across individuals. We cannot rule out the possibility that differences in vascular health are the consequence rather than the cause of differences in cognitive ability. Though the cross-sectional nature of our study cannot speak directly to the expansive body of literature on effects of mid-life blood pressure on late-life cognition (Obisesan et al., 2008; Mahinrad et al., 2020; Walker et al., 2019), the research can generate hypotheses to test in longitudinal datasets. The population-based adult-lifespan sample (18-88 years) used here is likely to be healthier, with lower variability in cardiovascular function, than samples used in other reports (Shafto et al., 2014). Cross-sectional data may also be limited by generational effects, including that recent decades have seen an increase in education and a decrease in population blood pressure. Nevertheless, ability discrepancy scores which normalize for crystalized intelligence may partly account for potential educational disadvantage endured by the oldest participants in this sample (as should our inclusion of education as a covariate of no interest).

We estimated a subset of potential vascular factors, based on a limited set of measures on a single visit. Our measures were relatively easy to acquire in practice on a large scale. The reactivity of the autonomic system to an event or stressor, which may be cognitive, emotional or physical in nature, e.g. phasic heart rate variability (Laborde et al., 2018), could prove more sensitive to the resting state heart rate variability estimates used here. It is possible that there are more than three Latent vascular factors, but we cannot test that here with six vascular measures (Watkins, 2018), and to do so would require a greater number of vascular measures to be collected. For example, future work could test whether cholesterol levels (Köbe et al., 2021) comprise an independent factor, or instead load on one or more of the three latent vascular factors identified here, or examine lifestyle factors such as physical activity, smoking and socio-economic status.

It may seem surprising that we found no evidence that medications related to vascular health contribute to ability discrepancy, or at least modulate the effects of our Latent vascular factors on ability discrepancy. This may be because the indirect effects of medication on ability discrepancy are mediated fully through their direct effects on Latent vascular factors. Alternatively, it could be that, while medications affect current Latent vascular factors, they may be given too late to prevent pre-medication levels of vascular factors like pulse pressure from already causing irreversible effects on cognitive ability (Stuhec et al., 2017). Or, that the effects of chronic stable medication are mitigated by homeostasis. Either way, future research could investigate the mechanisms through which Latent vascular factors mediate cognitive change, for example through damage to cerebral vessels, changes in brain perfusion or neuroinflammation, perhaps by direct manipulation of medications.

In summary, we show that vascular ageing has multifactorial relationships with cognitive ageing. Of the three latent vascular factors, an increase in the factor expressing pulse pressure was uniquely associated with the cognitive discrepancy score, and this relationship was stronger for older adults. We suggest that maintaining low pulse pressure may help to preserve cognitive function into old age.

## Data Availability

All data analysed in the present study can be requested from the Cambridge Centre for Ageing and Neuroscience (Cam-CAN).

## AUTHOR CONTRIBUTIONS

Designed research: D.L.O.K., R.N.A.H., K.A.T., L.K.T., J.B.R., Cam-CAN

Performed research: D.L.O.K., R.N.A.H., K.A.T.

Contributed unpublished reagents / analytical tools: N.W., R.K.

Analysed data: D.L.O.K., K.A.T.

Wrote the paper: D.L.O.K., K.A.T., R.N.A.H.

## FUNDERS

Cam-CAN was supported by the Biotechnology and Biological Sciences Research Council Grant BB/H008217/1.

D.L.O.K. was supported by a Doctoral Training Programme studentship awarded by the Biotechnology and Biological Sciences Research Council (BBSRC BB/M011194/1)

K.A.T. was supported by the Guarantors of Brain (G101149).

R.N.A.H. was supported by the UK Medical Research Council (SUAG/046 G101400).

J.B.R. was supported by the UK Medical Research Council (SUAG/051 G101400) and NIHR Cambridge Biomedical Research Centre (BRC-1215-20014; The views expressed are those of the authors and not necessarily those of the NIHR or the Department of Health and Social Care) and Wellcome Trust (220258).

We thank the Cam-CAN respondents and their primary care teams in Cambridge for their participation in this study, and colleagues at the MRC Cognition and Brain Sciences Unit MEG and MRI facilities for their assistance. Further information about the Cam-CAN corporate authorship membership can be found at http://www.cam-can.com/publications/Cam-CAN_Corporate_Author.html (**list #13**).

## Notes

### Competing Interest Statement

All authors have no conflicts of interest. Unrelated to this, there are several disclosures:
Cam-CAN was supported by the Biotechnology and Biological Sciences Research Council Grant BB/H008217/1.
D.L.O.K. was supported by a Doctoral Training Programme studentship awarded by the Biotechnology and Biological Sciences Research Council (BBSRC BB/M011194/1)
K.A.T. was supported by the Guarantors of Brain (G101149).
R.N.A.H. was supported by the UK Medical Research Council (SUAG/046 G101400).
J.B.R. was supported by the UK Medical Research Council (SUAG/051 G101400) and NIHR Cambridge Biomedical Research Centre (BRC-1215-20014; The views expressed are those of the authors and not necessarily those of the NIHR or the Department of Health and Social Care) and Wellcome Trust (220258). J.B.R. serves as an associate editor to Brain and is 518 a non-remunerated trustee of the Guarantors of Brain, Darwin College and the PSP Association (UK). He provides consultancy to Asceneuron, Biogen, UCB and has research grants from AZ-Medimmune, Janssen, and Lilly as industry partners in the Dementias Platform UK.
We thank the Cam-CAN respondents and their primary care teams in Cambridge for their participation in this study, and colleagues at the MRC Cognition and Brain Sciences Unit MEG and MRI facilities for their assistance. Further information about the Cam-CAN corporate authorship membership can be found at http://www.cam-can.com/publications/Cam-CAN_Corporate_Author.html (list #13).

### Author Declarations

Ethical approval was obtained from the Cambridgeshire 2 (now East of England Cambridge Central) Research Ethics Committee.

