## Supplementary Material for "Distinct components of cardiovascular health are linked with age-related differences in cognitive abilities"

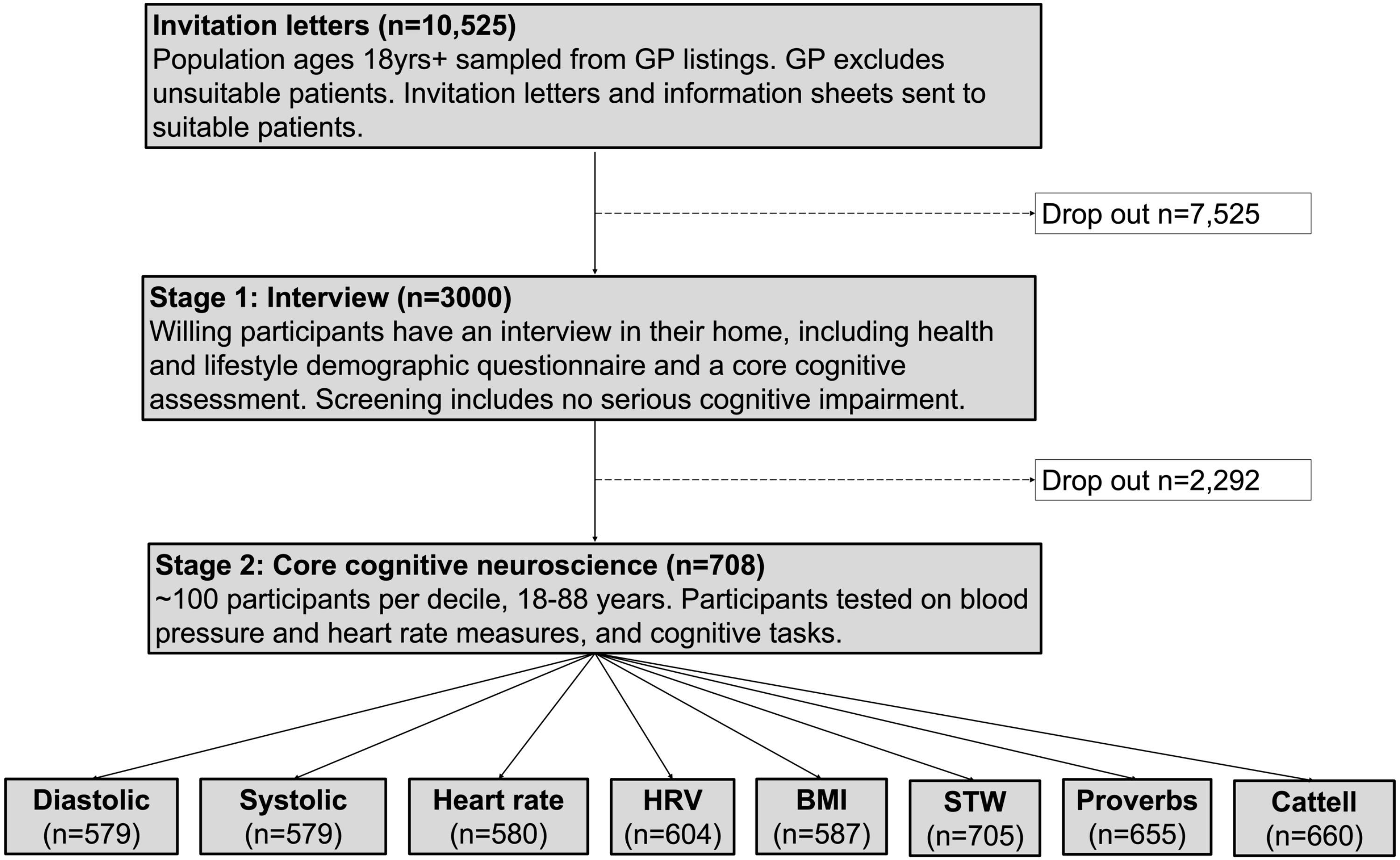


**Supplementary Figure 1**. Flowchart of participant recruitment in the Cam-CAN cohort, adapted from Shafto et al., 2014. Abbreviations: HRV, heart rate variability, recorded at low and high frequencies; STW, spot the word.

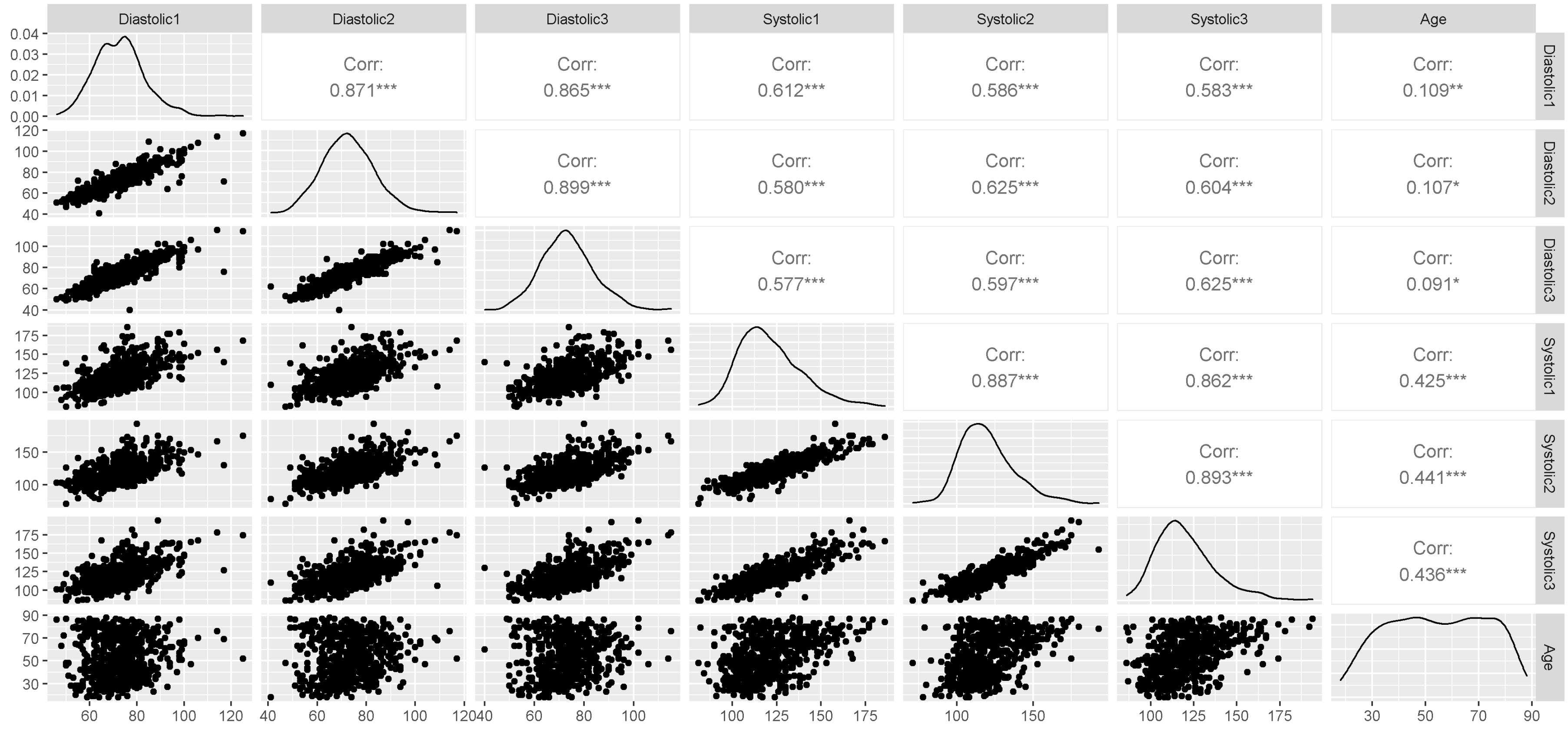


**Supplementary Figure 2**. Scatter plots (lower left), distributions (leading diagonal) and Pearson correlations (upper right) for age and diastolic and systolic blood pressure, across the first (n=578), second (n=579) and third (n=577) blood pressure recordings. Hypertension is clinically diagnosed when diastolic >90 mmHg and systolic >140 mmHg, or systolic >150mmHg for individuals aged over 80 (NICE, 2021). Stars indicate increasing significance on the correlations: ***, p<0.001; **, p<0.01; *, p<0.05.
Abbreviation: Corr, correlation coefficient.

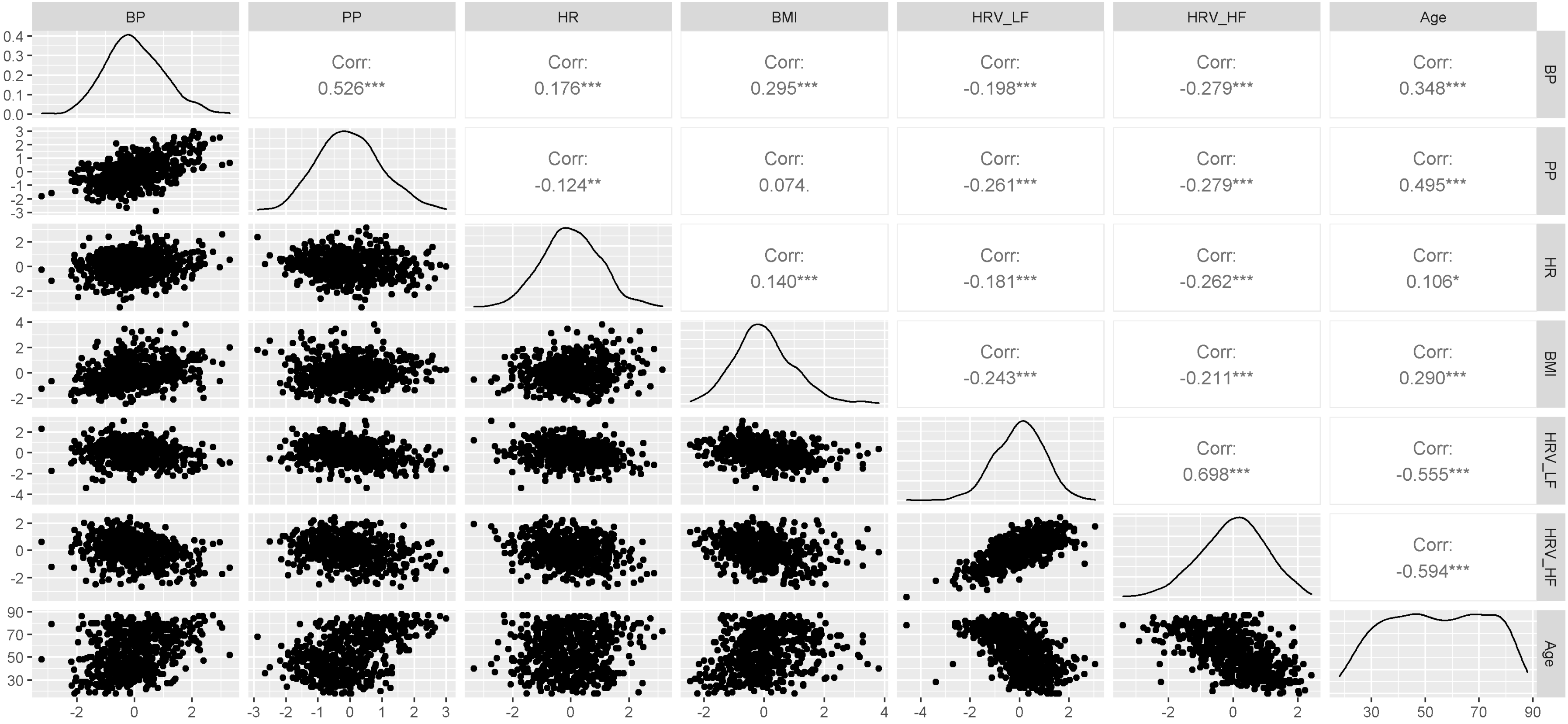


**Supplementary Figure 3**. Scatter plots (lower left), distributions (leading diagonal) and Pearson correlations (upper right) for vascular factors and age. Stars indicate increasing significance on the correlations: ***, p<0.001; **, p<0.01; *, p<0.05.
Abbreviations: BMI, body mass index; BP, total blood pressure; Corr, correlation coefficient; HR, heart rate; HRV_HF, heart rate variability at high frequency; HRV_LF, heart rate variability at low frequency; PP, pulse pressure.

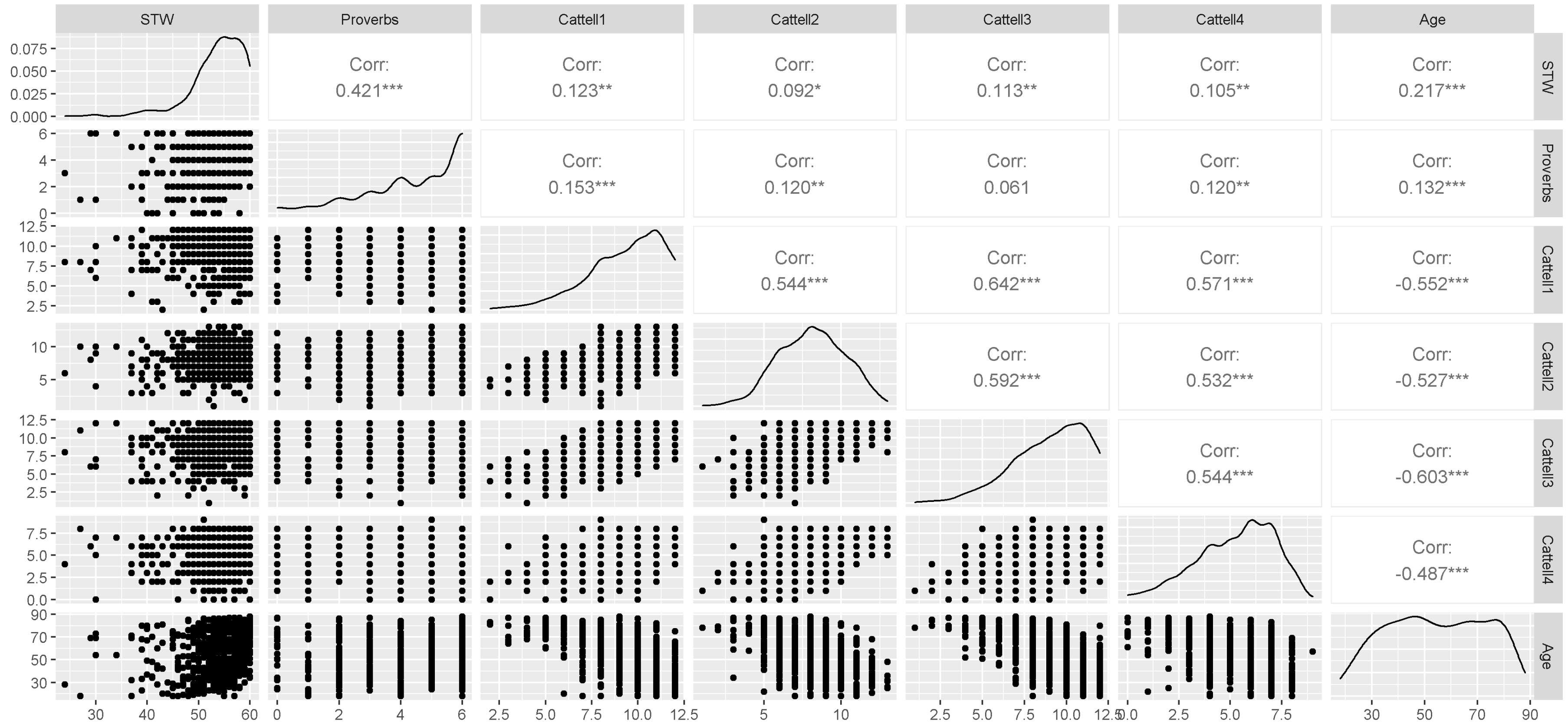


**Supplementary Figure 4**. Scatter plots (lower left), distributions (leading diagonal) and Pearson correlations (upper right) for cognitive observed variables and age. Stars indicate increasing significance on the correlations: ***, p<0.001; **, p<0.01; *, p<0.05. Abbreviations: Cattell 1-4, sub-scores across the four Cattell tasks; Corr, correlation coefficient; STW, spot the word.

**Supplementary Table 1.** Loadings of observed variables onto the latent variables in the winning 3 factor EFA model of vascular health (n=668).

| Predictors | LVF1 | LVF2 | LVF3 |
|---|---|---|---|
| HRV HF | -0.03 | < -0.01 | 0.85 |
| HRV LF | 0.06 | -0.05 | 0.82 |
| BP | 0.98 | 0.04 | 0.02 |
| BMI | 0.30 | -0.15 | -0.22 |
| PP | 0.06 | 0.91 | -0.05 |
| HR | 0.28 | -0.37 | -0.29 |

Abbreviations: BMI, body mass index; BP, total blood pressure; HR, heart rate; HRV HF, heart rate variability at high frequency; HRV LF, heart rate variability at low frequency; LVF, latent vascular factor; PP, pulse pressure.

**Supplementary Table 2.** Results of Model 1 (n=655, DoF=646, residual standard error =0.84). Significant effects ($p<0.05$) are shown in bold.

| Predictors | Standard β | Standard Error | Confidence Intervals | p |
|---|---|---|---|---|
| LVF1 | <0.01 | 0.04 | -0.08 – 0.08 | 1.00 |
| LVF2 | 0.21 | 0.04 | 0.12 – 0.29 | **<0.001** |
| LVF3 | -0.34 | 0.04 | -0.42 – -0.26 | **<0.001** |
| Sex | 0.06 | 0.07 | -0.08 – 0.20 | 0.42 |
| Education | | | | |
| No qualifications tried | -0.14 | 0.66 | -1.43 – 1.16 | 0.84 |
| GCSEs / O-levels | -0.62 | 0.65 | -1.91 – 0.66 | 0.34 |
| A-levels | -0.32 | 0.65 | -1.60 – 0.96 | 0.63 |
| Degree | -0.33 | 0.65 | -1.60 – 0.94 | 0.61 |

Abbreviations: LVF1-3, latent vascular factors.

**Supplementary Table 3.** Results of Model 2 (n=655, DoF=638, residual standard error =0.65). Significant effects ($p<0.05$) are shown in bold.

| Predictors | Standard β | Standard Error | Confidence Intervals | p |
|---|---|---|---|---|
| LVF1 | <0.01 | 0.03 | -0.06 – 0.07 | 0.90 |
| LVF2 | -0.02 | 0.04 | -0.10 – 0.05 | 0.56 |
| LVF3 | 0.02 | 0.04 | -0.06 – 0.09 | 0.65 |
| Age | 0.71 | 0.05 | 0.62 – 0.80 | **<0.001** |
| $Age^2$ | 0.03 | 0.04 | -0.04 – 0.11 | 0.37 |
| LVF1.Age | <0.01 | 0.03 | -0.07 – 0.06 | 0.94 |
| LVF1.$Age^2$ | -0.02 | 0.03 | -0.09 – 0.04 | 0.46 |
| LVF2.Age | <0.01 | 0.04 | -0.07 – 0.08 | 0.91 |
| LVF2.$Age^2$ | 0.08 | 0.04 | 0.01 – 0.15 | **0.03** |
| LVF3.Age | 0.01 | 0.04 | -0.06 – 0.08 | 0.73 |
| LVF3.$Age^2$ | <0.01 | 0.03 | -0.07 – 0.07 | 0.94 |
| Sex | 0.01 | 0.06 | -0.11 – 0.12 | 0.92 |
| Education | | | | |
| No qualifications tried | -0.64 | 0.51 | -1.65 – 0.37 | 0.21 |
| GCSEs / O-levels | -0.73 | 0.51 | -1.72 – 0.27 | 0.15 |
| A-levels | -0.50 | 0.50 | -1.49 – 0.49 | 0.32 |
| Degree | -0.37 | 0.50 | -1.36 – 0.61 | 0.46 |

Abbreviations: LVF1-3, latent vascular factors.

**Supplementary Table 4**. Results of model comparisons, using AIC and BIC, and the sum of squares derived from ANOVA comparisons.

| | Difference in AIC | Difference in BIC | Difference in Sum Of Squares |
|---|---|---|---|
| Model 1 vs 2 | 281.15 | 245.27 | 190.33 |
| Model 2 vs 3 | -22.34 | -237.61 | 35.26 |
| Model 2 vs 4 | -13.21 | -62.54 | 4.42 |
| Model 2 vs 5 | -18.10 | -71.92 | 2.97 |

Abbreviations: AIC, Akaike information criterion; BIC, Bayesian information criterion.

**Supplementary Table 5.** Results of Model 3 (n=655, DoF=590, residual standard error =0.61). Significant effects ($p<0.05$) are shown in bold. The adjusted p-value with Bonferroni corrections for this non-winning model would be $p<0.0008$.

| Predictors | Standard β | Standard Error | Confidence Intervals | p |
|---|---|---|---|---|
| LVF1 | -0.01 | 0.04 | -0.08 – 0.07 | 0.83 |
| LVF2 | 0.04 | 0.05 | -0.05 – 0.13 | 0.41 |
| LVF3 | -0.03 | 0.04 | -0.12 – 0.05 | 0.43 |
| Age | 0.64 | 0.05 | 0.54 – 0.75 | **<0.001** |
| $Age^2$ | -0.01 | 0.04 | -0.10 – 0.07 | 0.81 |
| LVF1.Age | -0.03 | 0.04 | -0.11 – 0.05 | 0.45 |
| LVF1.$Age^2$ | -0.05 | 0.04 | -0.12 – 0.03 | 0.22 |
| LVF2.Age | 0.05 | 0.05 | -0.04 – 0.14 | 0.27 |
| LVF2.$Age^2$ | 0.08 | 0.04 | -0.01 – 0.16 | 0.08 |
| LVF3.Age | -0.05 | 0.04 | -0.13 – 0.04 | 0.28 |
| LVF3.$Age^2$ | -0.02 | 0.04 | -0.10 – 0.06 | 0.64 |
| Anti-Hypertensives | 1.22 | 0.75 | -0.25 – 2.69 | 0.10 |
| Beta Blockers | 1.88 | 2.34 | -2.72 – 6.48 | 0.42 |
| Diuretics | -2.27 | 2.55 | -7.28 – 2.75 | 0.38 |
| Dyslipidemics | -0.66 | 0.5 | -1.64 – 0.32 | 0.19 |
| LVF1.Anti-Hypertensives | -1.46 | 1.28 | -3.98 – 1.06 | 0.26 |
| LVF1.Betablockers | -6.97 | 10.46 | -27.51 – 13.57 | 0.51 |
| LVF1.Diuretics | 7.00 | 3.16 | 0.79 – 13.20 | **0.03** |
| LVF1.Dyslipidemics | -0.83 | 0.52 | -1.85 – 0.20 | 0.11 |
| LVF2.Anti-Hypertensives | -1.04 | 0.69 | -2.40 – 0.31 | 0.13 |
| LVF2.Betablockers | 3.18 | 5.92 | -8.45 – 14.81 | 0.59 |
| LVF2.Diuretics | -3.32 | 3.92 | -11.02 – 4.39 | 0.40 |
| LVF2.Dyslipidemics | 1.16 | 0.61 | -0.05 – 2.37 | 0.06 |
| LVF3.Anti-Hypertensives | 0.29 | 1.01 | -1.69 – 2.28 | 0.77 |
| LVF3.Betablockers | -4.13 | 5.35 | -14.64 – 6.37 | 0.44 |
| LVF3.Diuretics | -0.7 | 2.87 | -6.33 – 4.94 | 0.81 |
| LVF3.Dyslipidemics | -0.66 | 1.15 | -2.91 – 1.59 | 0.57 |
| LVF1.Anti-Hypertensives.Age | 1.11 | 1.5 | -1.83 – 4.06 | 0.46 |
| LVF1.Anti-Hypertensives.$Age^2$ | -0.03 | 0.73 | -1.46 – 1.40 | 0.97 |
| LVF1.Betablockers.Age | 6.41 | 9.93 | -13.09 – 25.92 | 0.52 |
| LVF1.Betablockers.$Age^2$ | -1.21 | 3 | -7.10 – 4.69 | 0.69 |
| LVF1.Diuretics.Age | -7.05 | 3.41 | -13.75 – -0.35 | 0.04 |

| | | | | |
|---|---|---|---|---|
| LVF1.Diuretics.Age$^2$ | 2.39 | 1.31 | -0.18 – 4.95 | 0.07 |
| LVF1.Dyslipidemics.Age | 1.07 | 0.6 | -0.12 – 2.26 | 0.08 |
| LVF1.Dyslipidemics.Age$^2$ | -0.73 | 0.42 | -1.55 – 0.10 | 0.08 |
| LVF2.Anti-Hypertensives.Age | 1.25 | 0.84 | -0.40 – 2.89 | 0.14 |
| LVF2.Anti-Hypertensives.Age$^2$ | -0.92 | 0.48 | -1.87 – 0.02 | 0.06 |
| LVF2.Betablockers.Age | -3.56 | 5.53 | -14.43 – 7.30 | 0.52 |
| LVF2.Betablockers.Age$^2$ | 0.75 | 1.67 | -2.53 – 4.03 | 0.65 |
| LVF2.Diuretics.Age | 3.85 | 4.32 | -4.62 – 12.33 | 0.37 |
| LVF2.Diuretics.Age$^2$ | -1.38 | 1.87 | -5.04 – 2.29 | 0.46 |
| LVF2.Dyslipidemics.Age | -1.45 | 0.68 | -2.79 – -0.12 | **0.03** |
| LVF2.Dyslipidemics.Age$^2$ | 1.13 | 0.36 | 0.41 – 1.84 | **<0.01** |
| LVF3.Anti-Hypertensives.Age | -0.06 | 1.16 | -2.34 – 2.22 | 0.96 |
| LVF3.Anti-Hypertensives.Age$^2$ | 0.08 | 0.6 | -1.10 – 1.27 | 0.89 |
| LVF3.Betablockers.Age | 5.09 | 5.7 | -6.10 – 16.27 | 0.37 |
| LVF3.Betablockers.Age$^2$ | -2.77 | 2.52 | -7.71 – 2.17 | 0.27 |
| LVF3.Diuretics.Age | 1.12 | 3.09 | -4.95 – 7.19 | 0.72 |
| LVF3.Diuretics.Age$^2$ | -0.65 | 1.42 | -3.43 – 2.14 | 0.65 |
| LVF3.Dyslipidemics.Age | 0.62 | 1.27 | -1.87 – 3.12 | 0.62 |
| LVF3.Dyslipidemics.Age$^2$ | -0.25 | 0.63 | -1.49 – 0.99 | 0.69 |
| Anti-Hypertensives.Age | -1.07 | 0.91 | -2.85 – 0.71 | 0.24 |
| Anti-Hypertensives.Age$^2$ | 0.68 | 0.53 | -0.36 – 1.72 | 0.20 |
| Beta Blockers.Age | -0.86 | 2.44 | -5.66 – 3.94 | 0.72 |
| Beta Blockers.Age$^2$ | 0.15 | 1.21 | -2.22 – 2.53 | 0.90 |
| Diuretics.Age | 2.24 | 2.87 | -3.40 – 7.88 | 0.44 |
| Diuretics.Age$^2$ | -1.01 | 1.34 | -3.63 – 1.62 | 0.45 |
| Dyslipidemics.Age | 0.81 | 0.57 | -0.31 – 1.92 | 0.16 |
| Dyslipidemics.Age$^2$ | -0.66 | 0.4 | -1.46 – 0.13 | 0.10 |
| Sex | 0.04 | 0.06 | -0.08 – 0.16 | 0.46 |
| Education | | | | |
| No qualifications tried | -0.75 | 0.52 | -1.77 – 0.27 | 0.15 |
| GCSEs / O-levels | -0.81 | 0.51 | -1.81 – 0.19 | 0.11 |
| A-levels | -0.53 | 0.51 | -1.52 – 0.47 | 0.30 |
| Degree | -0.42 | 0.5 | -1.41 – 0.57 | 0.40 |

Abbreviations: LVF1-3, latent vascular factors.

**Supplementary Table 6.** Results of Model 4 (n=655, DoF=627, residual standard error =0.65). Significant effects ($p<0.05$) are shown in bold. The adjusted p-value with Bonferroni corrections for this non-winning model would be $p<0.002$.

| Predictors | Standard β | Standard Error | Confidence Intervals | p |
|---|---|---|---|---|
| LVF1 | -0.02 | 0.12 | -0.25 – 0.21 | 0.85 |
| LVF2 | -0.01 | 0.13 | -0.26 – 0.23 | 0.93 |
| LVF3 | 0.08 | 0.13 | -0.17 – 0.34 | 0.52 |
| Age | 0.58 | 0.15 | 0.29 – 0.87 | **<0.001** |
| $Age^2$ | 0.05 | 0.13 | -0.20 – 0.29 | 0.72 |
| LVF1.Age | -0.12 | 0.12 | -0.35 – 0.10 | 0.29 |
| LVF1.$Age^2$ | 0.06 | 0.11 | -0.15 – 0.28 | 0.55 |
| LVF2.Age | 0.04 | 0.13 | -0.22 – 0.31 | 0.74 |
| LVF2.$Age^2$ | 0.04 | 0.12 | -0.20 – 0.28 | 0.73 |
| LVF3.Age | 0.08 | 0.13 | -0.17 – 0.34 | 0.52 |
| LVF3.$Age^2$ | 0.09 | 0.12 | -0.14 – 0.33 | 0.44 |
| Sex | -0.06 | 0.08 | -0.22 – 0.11 | 0.50 |
| LVF1.Sex | 0.01 | 0.07 | -0.13 – 0.15 | 0.88 |
| LVF2.Sex | -0.01 | 0.08 | -0.16 – 0.15 | 0.92 |
| LVF3.Sex | -0.04 | 0.08 | -0.20 – 0.11 | 0.59 |
| Age.Sex | 0.10 | 0.09 | -0.09 – 0.28 | 0.30 |
| $Age^2$.Sex | -0.02 | 0.08 | -0.17 – 0.14 | 0.83 |
| LVF1.Age.Sex | 0.08 | 0.07 | -0.06 – 0.22 | 0.29 |
| LVF1.$Age^2$.Sex | -0.06 | 0.07 | -0.20 – 0.07 | 0.37 |
| LVF2.Age.Sex | -0.01 | 0.08 | -0.18 – 0.15 | 0.89 |
| LVF2.$Age^2$.Sex | 0.02 | 0.08 | -0.13 – 0.17 | 0.77 |
| LVF3.Age.Sex | -0.05 | 0.08 | -0.20 – 0.11 | 0.56 |
| LVF3.$Age^2$.Sex | -0.05 | 0.07 | -0.19 – 0.09 | 0.50 |
| Education | | | | |
| No qualifications tried | -0.58 | 0.53 | -1.63 – 0.47 | 0.28 |
| GCSEs / O-levels | -0.66 | 0.53 | -1.69 – 0.38 | 0.21 |
| A-levels | -0.44 | 0.52 | -1.47 – 0.59 | 0.40 |
| Degree | -0.31 | 0.52 | -1.33 – 0.72 | 0.56 |

Abbreviations: LVF1-3, latent vascular factors.

**Supplementary Table 7.** Results of Model 5 (n=655, DoF=626, residual standard error =0.64). Significant effects ($p<0.05$) are shown in bold. The adjusted p-value with Bonferroni corrections for this non-winning model would be $p<0.002$.

| Predictors | Standard β | Standard Error | Confidence Intervals | p |
|---|---|---|---|---|
| LVF1 | 0.01 | 0.05 | -0.08 – 0.10 | 0.83 |
| LVF2 | 0.01 | 0.05 | -0.09 – 0.11 | 0.90 |
| LVF3 | 0.01 | 0.05 | -0.08 – 0.11 | 0.80 |
| Age | 0.71 | 0.05 | 0.62 – 0.81 | **<0.001** |
| $Age^2$ | 0.02 | 0.04 | -0.07 – 0.10 | 0.66 |
| LVF1.Age | -0.01 | 0.05 | -0.11 – 0.09 | 0.90 |
| LVF1.$Age^2$ | -0.03 | 0.04 | -0.11 – 0.05 | 0.49 |
| LVF2.Age | 0.01 | 0.06 | -0.10 – 0.12 | 0.91 |
| LVF2.$Age^2$ | 0.09 | 0.04 | 0.01 – 0.18 | **0.04** |
| LVF3.Age | -0.01 | 0.05 | -0.10 – 0.09 | 0.88 |
| LVF3.$Age^2$ | 0.01 | 0.04 | -0.08 – 0.09 | 0.85 |
| LVF1.LVF2 | 0.01 | 0.05 | -0.08 – 0.10 | 0.81 |
| LVF1.LVF3 | 0.03 | 0.04 | -0.06 – 0.11 | 0.54 |
| LVF2.LVF3 | <0.01 | 0.06 | -0.11 – 0.11 | 0.96 |
| LVF1.LVF2.LVF3 | 0.03 | 0.04 | -0.06 – 0.11 | 0.56 |
| LVF1.LVF2.Age | -0.04 | 0.05 | -0.12 – 0.05 | 0.44 |
| LVF1.LVF2.$Age^2$ | <0.01 | 0.03 | -0.06 – 0.06 | 1.00 |
| LVF1.LVF3.Age | -0.04 | 0.05 | -0.13 – 0.05 | 0.39 |
| LVF1.LVF3.$Age^2$ | -0.05 | 0.04 | -0.13 – 0.03 | 0.26 |
| LVF2.LVF3.Age | 0.02 | 0.06 | -0.10 – 0.14 | 0.78 |
| LVF2.LVF3.$Age^2$ | 0.01 | 0.05 | -0.08 – 0.11 | 0.78 |
| LVF1.LVF2.LVF3.Age | -0.01 | 0.05 | -0.10 – 0.09 | 0.92 |
| LVF1.LVF2.LVF3.$Age^2$ | -0.01 | 0.03 | -0.07 – 0.05 | 0.76 |
| Sex | <0.01 | 0.06 | -0.12 – 0.12 | 0.99 |
| Education | | | | |
| No qualifications tried | -0.62 | 0.53 | -1.65 – 0.42 | 0.25 |
| GCSEs / O-levels | -0.7 | 0.52 | -1.73 – 0.32 | 0.18 |
| A-levels | -0.48 | 0.52 | -1.50 – 0.54 | 0.36 |
| Degree | -0.35 | 0.52 | -1.37 – 0.66 | 0.49 |

Abbreviations: LVF1-3, latent vascular factors.

SUPPLEMENTARY MATERIAL: SECTION B

### Introduction

The winning 3-factor EFA model (Figure 2) was saturated and therefore lacked absolute fit indices. The robustness of this model was investigated using Exploratory Structural Equation Modelling (ESEM) (Asparouhov et al., 2019). ESEM integrates confirmatory factor analysis and structural equation modelling to provide confirmatory tests of a priori factor structures. It allowed modelling of vascular and cognitive factors simultaneously. We hypothesised a winning model with three latent vascular factors (Chen et al., 2000; Goodman et al., 2005; Khader et al., 2011; Mayer-Davis et al., 2009; Tsvetanov et al., 2021b), and with two latent cognitive factors representing the distinct domains of fluid and crystallized intelligence (Borgeest et al., 2020; Cattell, 1943; McDonough et al., 2016). We further hypothesised that the three latent vascular factors in the ESEM model would structurally resemble and correlate highly with the corresponding latent vascular factors produced in the main EFA analysis (Figure 2). This would evidence that a 3-factor structure robustly and reliably captures the vascular variables (blood pressure, pulse pressure, BMI, heart rate and heart rate variability). This could allow advances in the design of studies to understand the links between vascular health and cognitive ageing.

### Methods

Participants with <2 observations in either vascular or cognitive variables were excluded (n=655). Confirmatory Factor Analysis produced latent factors, as in the main analysis (Figure 1). Then all vascular and cognitive variables were input to ESEM models, estimated with the Psych package (Revelle, 2017). The ESEM models produced latent vascular factors and LCFs using only participants with complete data across all variables for each domain (Latent vascular factors n=516, latent cognitive factors n=636). Iterative combinations of 1-4 latent vascular factors and 1-4 LCFs were explored. Model fit was evaluated according to the Bayesian Information Criterion (BIC), adjusted for sample size. From the winning model, subject scores were extracted that represent how each participant's observed data loads onto the latent vascular factors. The latent vascular factors produced in the ESEM model were correlated with those produced in EFA.

### Results

The winning ESEM model has 3 vascular and 2 cognitive factors, indicated as "3_2" in Supplementary Figure 5. The latent vascular factors produced in ESEM correlated highly those produced in EFA ($r>0.99$, $p<0.001$, in all instances; Supplementary Figure 6).

### **Discussion**

The vascular structure is robustly and reliably produced in EFA and ESEM data-driven models. This provides strong support for the initial EFA model, in the absence of absolute fit indices. The EFA vascular structure could be further confirmed in future studies by expanding the model with additional variables, such as heart rate variability reactivity, orthostatic intolerance or pulse wave velocity. It would also be possible to extend the ESEM analysis by calculating the ability discrepancy score from the latent cognitive factors, which resemble crystallized and fluid intelligence (McDonough et al., 2016). The discrepancy could then be related to latent vascular factors in multiple linear regression, as in the main analysis of the present study. However, this was not implemented because to model the vascular and cognitive factors simultaneously in ESEM, and to then relate them again in regression, is circular. In Summary, the multifactorial vascular structure is robust and will inform the design of future studies on vascular and cognitive ageing.

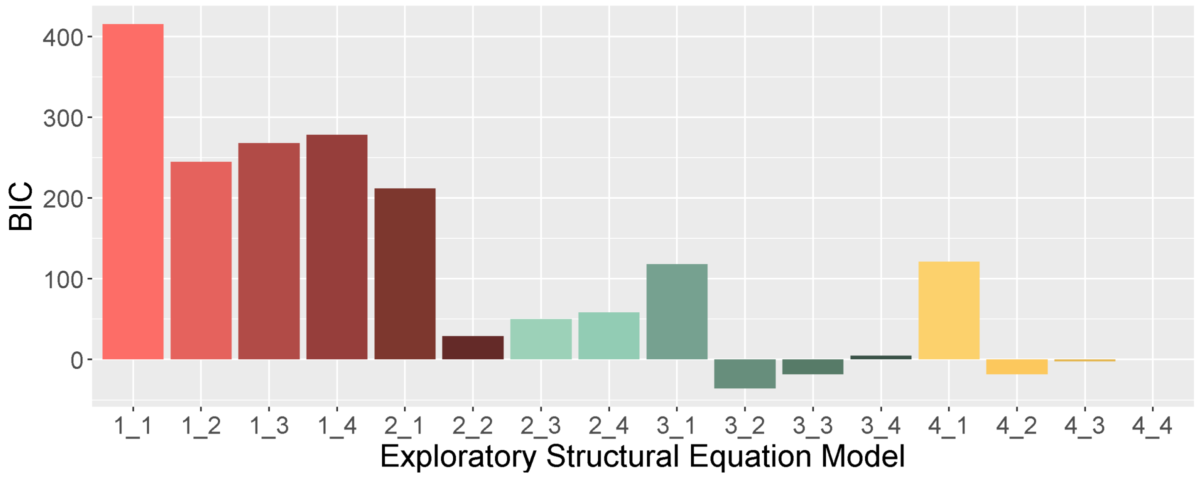


**Supplementary Figure 5.** A comparison of model fit for Exploratory Structural Equation Models. Individual models are each coloured and labelled on the x-axis with two digits, separated by an underscore, of which the first represents the number of vascular and the second the number of cognitive latent variables within a given model. Fit is measured by the Bayesian Information Criterion (BIC). Model “3_2” has the best overall fit and was selected for further examination.

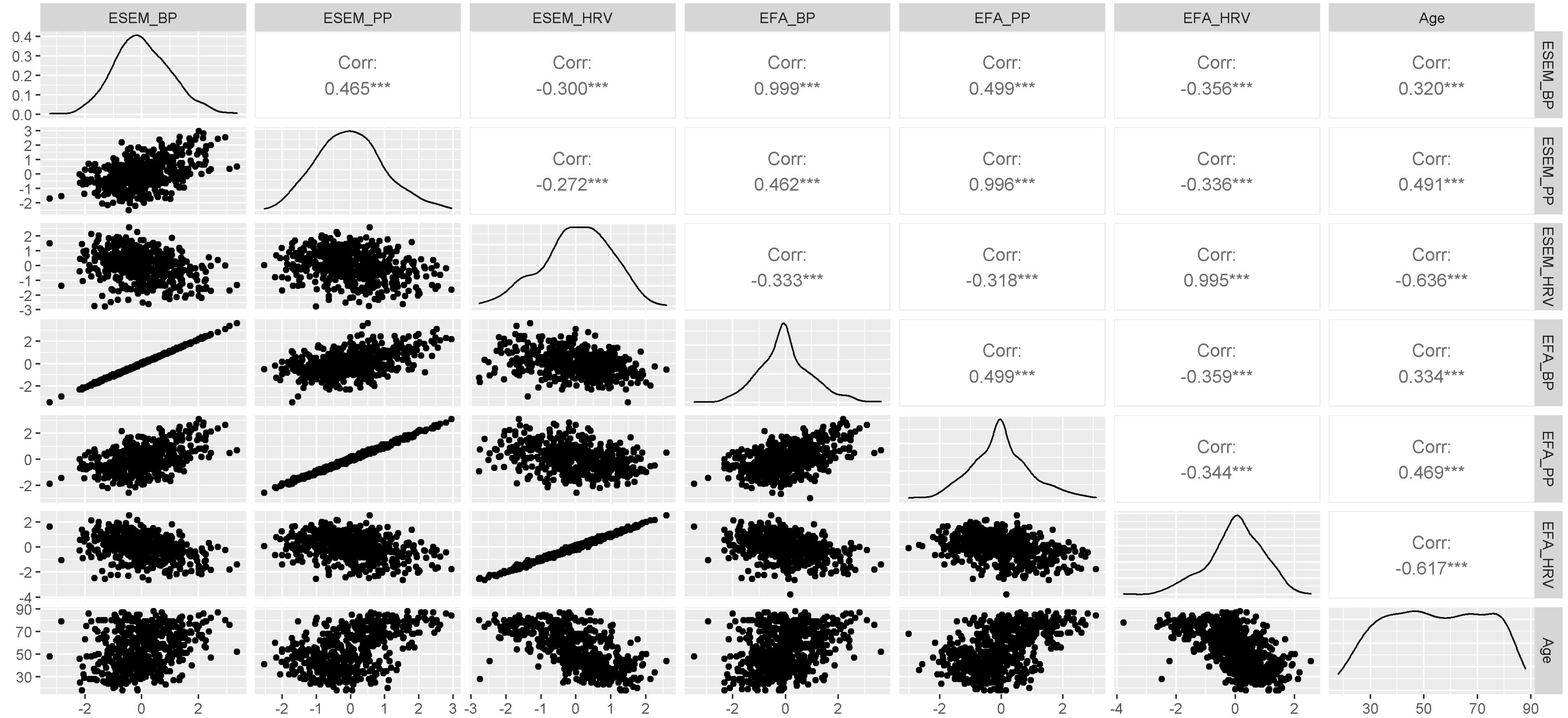


**Supplementary Figure 6**. Scatter plots (lower left), distributions (leading diagonal) and Pearson correlations (upper right) for Latent vascular factors produced in ESEM and EFA, and age. Latent vascular factors are named according to the observed variable(s) predominantly expressed: blood pressure (BP), pulse pressure (PP) and heart rate variability (HRV). Stars indicate increasing significance on the correlations: ***, p<0.001; **, p<0.01; *, p<0.05.

Abbreviations: Corr, correlation coefficient; EFA, Exploratory Factor Analysis; ESEM, Exploratory Structural Equation Modelling.
